# Seroprevalence of SARS-CoV-2 Antibodies in Seattle, Washington—October 2019–April 2020

**DOI:** 10.1101/2020.12.07.20244103

**Authors:** Denise J. McCulloch, Michael L. Jackson, James P. Hughes, Sandra Lester, Lisa Mills, Brandi Freeman, Mohammad Ata Ut Rasheed, Natalie J. Thornburg, Helen Y. Chu

## Abstract

Estimating prevalence of SARS-CoV-2 antibodies is important to determine disease burden. We tested residual samples from 763 Seattle-area adults for SARS-CoV-2 antibodies. Prevalence rose from 0% to 1.2% between October 2019–April 2020, suggesting a small percentage of this metropolitan-area cohort had been infected with SARS-CoV-2 at that time.

## Introduction

Estimating the cumulative incidence of SARS-CoV-2 infection is critical for understanding the progression of local COVID-19 epidemics. Early in the course of the SARS-CoV-2 pandemic, most data on COVID-19 came from hospitalized patients due to limitations in the availability of SARS-COV-2 tests. The lack of widely available testing for milder illnesses left most public health officials with an incomplete picture of incidence in the community, including cumulative incidence of infection and potential progress toward herd immunity. Serologic surveys for SARS-CoV-2 antibodies can help fill this knowledge gap.^1,2^

The greater Seattle area of western Washington State was the site of the first detected COVID-19 case in the United States. Detection of this case on 20 January 2020 was followed by cryptic SARS-CoV-2 transmission until discovery of additional COVD-19 cases and the first known COVID-19 death on 29 February 2020.^3^ This was followed by the declaration of a state of emergency and multiple social distancing interventions aimed at curbing spread of SARS-CoV-2. To evaluate the effects of local control measures and inform future interventions, we estimated the prevalence of SARS-CoV-2 infection in the greater Seattle area through early April 2020.

## Methods

Residual sera were obtained from the virology laboratory at the University of Washington Medical Center. They were collected from inpatients and outpatients >18 years who underwent routine screening for hepatitis viruses. Up to 114 sera were collected on each collection date. Residual sera were collected one day per month during October through January, were not collected in February, and then were collected weekly beginning in March. Residual sera after routine hepatitis screening were collected by a laboratory technician in Dr. Helen Chu’s laboratory. On the day of collection, sera were transported from the clinic to the Chu lab in a cooler with ice packs, aliquoted and immediately frozen at −20°C into three aliquots of 500-750 ul.

Serum samples were shipped frozen overnight on dry ice on Wednesday, March 25 and Monday, April 13 to the Centers for Disease Control and Prevention (CDC) for testing and were received in good condition. Sera were diluted at 1:100 and pan-IgG secondary antibody, which can detect IgM, IgG, and/or IgA was used. Samples were tested with a SARS-CoV-2-specific-enzyme linked immunosorbent assay (ELISA) using the prefusion-stabilized form of the spike protein ^11^. Samples were considered seropositive if the anti-SARS-CoV-2 optical density (OD) spike was equal to or greater than a cutoff of 0.4. This cutoff produced a sensitivity of 96% and a specificity of 99.3% ^11^. After initial testing, sera were stored at four degrees for less than 2 days, and all positive samples underwent repeat testing with the same assay (to reduce the possibility of false positive results), and specimens were not considered positive unless they tested positive both times.

Data were analyzed in SAS 9.4 (Cary, NC). This study was approved by the Institutional Review Board of the University of Washington (STUDY #00006181).

## Results

Samples from 770 participants were sent for testing and 766 were of sufficient volume to perform ELISAs. Demographic data were available for 75% of samples. The median age of participants was 45 years (interquartile range, 32.5 - 60), and 50.8% were female.

All 261 samples from 2019 had OD values below the cutoff for SARS-CoV-2 antibodies (Table 1). Among 87 samples from January 8, 2020, (n = 3) 3.4% had OD values just above the cutoff on initial testing but below the OD cutoff upon repeat testing. Similarly, of 413 samples collected after March 1^st^, two sera samples collected on March 13, one collected on March 25, 2020, and one collected on April 1 had OD values just above the cutoff on initial testing but below the cutoff on repeat testing. These 3 samples from 2020 were also considered to be negative.

**Table 1.** Proportion of specimens testing positive for SARS-CoV-2 antibodies via ELISA assay by week of specimen collection.

| Week of sample collection | Positive<br>n=5 | Negative<br>n=761 | Total<br>n=766 | Estimated point prevalence (95% CI) |
| --- | --- | --- | --- | --- |
| October 2, 2019 | 0 (0%) | 87 (100%) | 87 |  |
| November 5, 2019 | 0 (0%) | 89 (100%) | 89 |  |
| December 6, 2019 | 0 (0%) | 87 (100%) | 87 |  |
| January 8, 2020 | 0 (0%) | 90 (100%) | 90 |  |
| March 5, 2020 | 0 (0%) | 5 (100%) | 5 |  |
| March 13, 2020 | 0 (0%) | 114 (100%) | 114 |  |
| March 18, 2020 | 1 (0.92%) | 108 (99.1%) | 109 | 0.92% (0.02 - 5.0%) |
| March 25, 2020 | 3 (3.33%) | 87 (96.7%) | 90 | 3.33% (0.7 - 9.4%) |
| April 1, 2020 | 1 (1.05%) | 94 (98.95%) | 95 | 1.05% (0.03 - 5.7%) |
| Total for March-April samples | 5 (1.21%) | 408 (98.79%) | 413 | 1.21% (0.4 - 2.8%) |
| Total | 5 (0.7%) | 761 (99.3%) | 766 |  |

The first confirmed positive serum was identified from one of the 107 samples collected on March 25. Four additional positive samples were identified in March and April (Table 1). The mean OD for the 5 confirmed positive samples was 1.41 (standard deviation, 0.58). The estimated prevalence of SARS-CoV-2 antibodies in our study population during March 5–April 1, 2020 was 1.2%.

## Discussion and Conclusions

The population prevalence of antibodies to SARS-CoV-2 in the greater Seattle area in March and early April 2020 was 1.2%. This is comparable to the seroprevalence reported in Seattle-area children around the same time period.^12^ Considering the epidemic may have started earlier in Washington state than other regions of the country, 1.2% is low compared to cities with large epidemics such as Lombardy or Madrid,^1,13^ supporting the idea that mitigation measures taken in WA successfully limited the early SARS-CoV-2 outbreak.

As of April 1, 2020, there were 2,468 confirmed cases of COVID-19 identified in King County ^14^, with a population of 2.253 million people. Therefore, at the start of April, there were 109.5 confirmed cases per 100,000 people in King County, WA. A seroprevalence of 1.2% corresponds to total infections being approximately 11 times greater than the number of confirmed cases in King County, though the sampling of participants in this study may make it unreliable to extrapolate to the general population. This ratio is comparable to the findings from New York, where 9% of infections were estimated to have been diagnosed during March and April.^8^

In contrast, a serosurvey in Santa Clara, CA, estimated that seroprevalence of SARS-CoV-2 antibodies was 50 to 85 times higher than the number of cases detected by diagnostic viral RT-PCR, and 43 times greater in Los Angeles, CA ^9^. The authors of the Santa Clara study acknowledged that recruitment via social media may have selected for individuals with a recent COVID-like illness seeking confirmation that an illness they had experienced could have been COVID-19. Furthermore, both the Santa Clara study used a point-of-care lateral flow assay, and concerns about the specificity of the assay used in this and other studies raise the possibility of false positives inflating the number of positive antibody test results^15^.

ELISA has been shown to have greater specificity for the detection of SARS-CoV-2 antibodies compared to lateral flow antibody assays^15^, thereby reducing the likelihood of false positives in our study. Additionally, all borderline or positive antibody results in our study underwent repeat testing to confirm positive results, further reducing the likelihood of our obtaining a false positive result.

Our approach has some advantages over serosurveys conducted in other regions. First, our study included samples from October–December, 2019, a time when COVID-19 was not known to be circulating. The absence of positive test results from samples during this time period reinforces that the positive results obtained from later samples in this study were unlikely to be false positives. Also, the collection of samples over several months allowed us to assess changes in population seroprevalence over time.

This study has several limitations. First, the relatively small sample size, involving hundreds rather than thousands of samples, limits the accuracy of our estimates. Second, the use of residual samples from patients tested for hepatitis selected for a group of patients who had contact with the healthcare system and may not be representative of the Seattle population as a whole. The generalizability of our findings might therefore be limited. Third, due to the use of de-identified samples, we are not able to describe the study population in detail in order to understand the representativeness of the sample. Fourth, we were unable to collect specimens in February, and SARS-CoV-2 may have been circulating in that month. Fifth, with stay at home orders in March and April, the sample population may have varied over time. Individuals who were seeking care in March and April may have had more serious conditions than those seeking care October-January. Lastly the median age of our study population is relatively young.

In spite of these limitations, this study contributes important data to the limited information we have thus far on the seroprevalence of antibodies to SARS-CoV-2. First, SARS-CoV-2 seroprevalence increased from zero before March 18 to 1.2% in late March/early April, consistent with the time frame of increasing confirmed COVID-19 cases in the Seattle-area at that time, corroborating the known time-frame of community spread of the virus^16^. Second, the low percentage of Seattle-area adults with serologic evidence of prior SARS-CoV-2 infection indicates that the vast majority of the local population may remain susceptible to COVID-19. This has important implications for public health as the pandemic evolves.

Future studies of population prevalence using specimens collected from a statistically representative cross sectional cohort, over time and across diverse geographic locations, are needed to help better characterize the true incidence and public health impact of COVID-19, thereby enabling more accurate estimations of infections, and mortality rates.

## Data Availability

The data analyzed during the current study are available from the corresponding author on reasonable request.

## Disclaimer

The findings and conclusions in this report are those of the author(s) and do not necessarily represent the official position of the Centers for Disease Control and Prevention. Names of specific vendors, manufacturers, or products are included for public health and informational purposes; inclusion does not imply endorsement of the vendors, manufacturers, or products by the Centers for Disease Control and Prevention or the US Department of Health and Human Services.

## References

1. Percivalle E, Cambie G, Cassaniti I, et al. Prevalence of SARS-CoV-2 specific neutralising antibodies in blood donors from the Lodi Red Zone in Lombardy, Italy, as at 06 April 2020. Euro Surveill. 2020;25(24).

2. Stringhini S, Wisniak A, Piumatti G, et al. Seroprevalence of anti-SARS-CoV-2 IgG antibodies in Geneva, Switzerland (SEROCoV-POP): a population-based study. Lancet. 2020.

3. COVID-19 Resources and Information | Governor Jay Inslee. 2020; https://www.governor.wa.gov/issues/issues/covid-19-resources. Accessed 3 June 2020.

4. Gudbjartsson DF, Helgason A, Jonsson H, et al. Spread of SARS-CoV-2 in the Icelandic Population. N Engl J Med. 2020;382(24):2302–2315.

5. Carvalho T. COVID-19 Research in Brief: 11 April to 17 April, 2020. Nat Med. 2020.

6. Ng D, Goldgof G, Shy B, et al. SARS-CoV-2 seroprevalence and neutralizing activity in donor and patient blood from the San Francisco Bay Area. medRxiv. 2020:2020.2005.2019.20107482.

7. Zou J, Bretin A, Gewirtz A. Antibodies to SARS/CoV-2 in arbitrarily-selected Atlanta residents. medRxiv. 2020:2020.2005.2001.20087478.

8. Rosenberg ES, Tesoriero JM, Rosenthal EM, et al. Cumulative incidence and diagnosis of SARS-CoV-2 infection in New York. medRxiv. 2020:2020.2005.2025.20113050.

9. Bendavid E, Mulaney B, Sood N, et al. COVID-19 Antibody Seroprevalence in Santa Clara County, California. medRxiv. 2020:2020.2004.2014.20062463.

10. Sood N, Simon P, Ebner P, et al. Seroprevalence of SARS-CoV-2-Specific Antibodies Among Adults in Los Angeles County, California, on April 10-11, 2020. JAMA. 2020.

11. Freeman B, Lester S, Mills L, et al. Validation of a SARS-CoV-2 spike protein ELISA for use in contact investigations and sero-surveillance. bioRxiv. 2020:2020.2004.2024.057323.

12. Dingens AS, Crawford KHD, Adler A, et al. Seroprevalence of SARS-CoV-2 among children visiting a hospital during the initial Seattle outbreak. medRxiv. 2020:2020.2005.2026.20114124.

13. Pollan M, Perez-Gomez B, Pastor-Barriuso R, et al. Prevalence of SARS-CoV-2 in Spain (ENE-COVID): a nationwide, population-based seroepidemiological study. Lancet. 2020.

14. Coronavirus daily news updates, April 1: What to know today about COVID-19 in the Seattle area, Washington state and the nation. The Seattle Times. April 1, 2020, 2020.

15. Adams ER, Anand R, Andersson MI, et al. Evaluation of antibody testing for SARS-Cov-2 using ELISA and lateral flow immunoassays. medRxiv. 2020:2020.2004.2015.20066407.

16. Washington State Department of Health. COVID-19 Data Dashboard. https://www.doh.wa.gov/Emergencies/NovelCoronavirusOutbreak2020COVID19/DataDashboard.

